# Breast milk exposure is associated with cortical maturation in preterm infants

**DOI:** 10.1101/2022.01.04.22268723

**Authors:** Gemma Sullivan, Kadi Vaher, Manuel Blesa, Paola Galdi, David Q. Stoye, Alan J. Quigley, Michael J. Thrippleton, Mark E. Bastin, James P. Boardman

**Affiliations:** MRC Centre for Reproductive Health, Queen’s Medical Research Institute, University of Edinburgh, Edinburgh, EH16 4TJ; Department of Radiology, Royal Hospital for Children and Young People, Edinburgh, EH16 4TJ; Edinburgh Imaging, University of Edinburgh, Edinburgh, EH16 4TJ; Centre for Clinical Brain Sciences, Chancellor’s Building, University of Edinburgh, Edinburgh, EH16 4SB

## Abstract

**Objective:** Breast milk exposure is associated with improved neurocognitive outcomes following preterm birth but the neural substrates linking nutrition with outcome are uncertain. By combining nutritional data with brain MRI, we tested the hypothesis that high versus low breast milk exposure in preterm infants during neonatal care results in a cortical morphology that more closely resembles that of infants born at term.

**Methods:** We studied 135 preterm (mean gestational age 30^+2^ weeks, range 22^+1^ to 32^+6^) and 77 term-born infants (mean gestational age 39^+4^ weeks, range 36^+3^ to 42^+1^). Nutritional data was collected from birth until hospital discharge to identify the proportion of days preterm infants received exclusive breast milk. Structural and diffusion MRI were performed at term-equivalent age. Cortical indices (volume, thickness, surface area, gyrification index, sulcal depth, curvature) and water diffusion parameters (fractional anisotropy, mean diffusivity, radial diffusivity, axial diffusivity, neurite density index, orientation dispersion index) were compared between preterm infants who received exclusive breast milk for <75% of inpatient days (*n*=68), preterm infants who received exclusive breast milk for ≥75% of inpatient days (*n*=67) and term-born controls (*n*=77).

**Results:** High breast milk exposure was associated with reduced cortical gray matter volume (*d*=0.47, *p*=0.014), thickness (*d*=0.42, *p*=0.039) and radial diffusivity (*d*=0.38, *p*=0.039), and increased fractional anisotropy (*d*=0.38, *p*=0.037) after adjustment for age at MRI.

**Interpretation:** High versus low breast milk exposure in the weeks following preterm birth is associated with a cortical imaging phenotype that more closely resembles the brain morphology of healthy infants born at term.

## Introduction

Breast milk feeding following preterm birth is associated with improved neurocognitive outcomes, academic performance and higher IQ scores^1-3^ but the neural substrates linking nutrition with outcome are uncertain. Neonatal MRI studies have shown that breast milk exposure in the weeks after preterm birth enhances brain growth and white matter microstructure^4-6^ but effects on early cortical development have not yet been studied. Breast milk duration and exclusivity are associated with improved measures of cortical maturation in term-born children^7, 8^ and adolescents^9^ but the extent to which this can be extrapolated to preterm infants is unknown because preterm birth itself affects cortical development, and nutrition and other confounding exposures through infancy and childhood can influence cortical growth. Resolving this uncertainty has important implications for neuroprotection strategies based on nutrition in the neonatal intensive care unit (NICU).

The third trimester of pregnancy is a period of rapid cortical development that involves multiple dynamic cellular processes and the establishment of neural circuits.^10^ During this critical period the cortex is therefore vulnerable to injury or dysmaturation due to early removal from the placental circulation and intrauterine environment, and exposure to environmental adversities associated with preterm birth; indeed, altered cortical development is a recognised feature of the encephalopathy of prematurity.^11^ Preterm infants at term-equivalent age have altered cortical growth, folding and microstructure when compared to term-born controls.^12, 13^ Cortical alterations persist into childhood^14, 15^ and adolescence^16^ and have been associated with impaired motor, cognitive and behavioural outcomes,^17, 18^ which focuses attention on the cortex as a neural substrate for neurodevelopmental impairment following preterm birth.

Optimal macronutrient intake after preterm birth can modify head growth and improve neurodevelopmental outcomes,^19, 20^ but studies have tended to focus on parenteral nutrition in the immediate postnatal period and relatively less attention has been given to the impact of nutrition in the weeks after the establishment of enteral feeding but before discharge from NICU which can be two to three months during the critical window of development. Exclusive breast milk feeding is associated with reduced growth velocity in preterm infants when compared to those fed formula milk and this can influence clinical decision-making regarding supplementation of maternal milk with formula. However, large cohort studies have shown that breast milk feeding is associated with reduced risk of neurodevelopmental impairment in childhood despite the higher risk of sub-optimal weight gain during the neonatal period.^21^

In this study, we combined nutritional data with brain MRI to test the hypothesis that high compared to low breast milk exposure in the weeks following preterm birth enhances early cortical development, resulting in a cortical architecture that more closely resembles that of healthy infants born at full term. We investigated the influence of breast milk exposure during neonatal care on a comprehensive set of cortical features derived from structural and diffusion MRI: volume, gyrification index, thickness, sulcal depth, curvature, surface area, fractional anisotropy (FA), mean diffusivity (MD), axial diffusivity (AD), radial diffusivity (RD), neurite density index (NDI) and orientation dispersion index (ODI).

## Materials and methods

### Participants

Participants were 135 preterm infants born before 32 weeks’ gestation and 77 term-born infants, delivered at the Royal Infirmary of Edinburgh, UK and recruited to the Theirworld Edinburgh Birth Cohort (TEBC)^**22**^ between February 2017 and December 2020. Ethical approval was obtained from the UK National Research Ethics Service and parents provided written informed consent (South East Scotland Research Ethic Committee 16/SS/0154) in accordance with the declaration of Helsinki. Exclusion criteria were major congenital malformation, chromosomal abnormality, congenital infection, cystic periventricular leukomalacia, haemorrhagic parenchymal infarction and post-haemorrhagic ventricular dilatation.

### Breast milk exposure

Daily nutritional intake for preterm infants was collected from birth until discharge from the neonatal unit using electronic patient records. Breast milk exposure was defined as the proportion (%) of inpatient days infants received exclusive breast milk feeds, which included both maternal and/or donor expressed breast milk. Infants were categorised into two groups based on breast milk exposure: High breast milk exposure was defined as exclusive breast milk feeds for ≥75% of inpatient days and low breast milk exposure was defined as exclusive breast milk feeds for <75% of inpatient days.

The nutritional management of all preterm participants included the administration of standard parenteral nutrition on admission to the neonatal unit (Scottish Neonatal Parenteral Nutrition 2.4 g/100ml, ITH Pharma, London) commencing at 100 ml/kg/day if birthweight <1000 g and 75 ml/kg/day if birthweight 1000-1500 g, incrementing daily to reach a total of 150 ml/kg/day. Fat and fat-soluble vitamins were provided using SMOF lipid 20% emulsion commenced at 1 g/kg/day within 24 hours of admission, increasing to 2 g/kg/day on day 2 and 3 g/kg/day on day 3. Expression of breast milk was supported immediately following delivery and colostrum was given as soon as it was available. Enteral feeds were commenced at 12 ml/kg/day on day 1. For low risk infants ≥28 weeks and ≥1000 g, feeds were subsequently incremented by 30 ml/kg/day. For infants <28 weeks or <1000 g, feeds were incremented by 18 ml/kg/day. For infants at any gestation with additional risk factors for necrotising enterocolitis (intra-uterine growth restriction, absent end-diastolic flow on umbilical artery doppler, monochorionic twins), feeds continued at 12 ml/kg/day for the first 48 hours with subsequent daily increments of 18 ml/kg/day. If there was insufficient maternal milk by 48 hours of age, donor expressed milk was given to supplement maternal supply for infants with birthweight <1500 g. Parenteral nutrition was stopped once infants reached an enteral volume of 120 ml/kg/day and feeds continued to increment to a maximum volume of 180-200 ml/kg/day. Human milk fortifier (Cow & Gate Nutriprem Human Milk Fortifier, Nutricia) was added to breast milk at 2-3 weeks of age. If there was insufficient maternal breast milk to meet infant requirement beyond 30 weeks corrected age, donor expressed milk was replaced with preterm formula. All infants received multivitamins from day 7 of life and iron supplementation from day 42. Family integrated care was encouraged throughout neonatal admission with opportunities for skin-to-skin contact and support for parents to perform routine care and nasogastric feeding.

### Image acquisition

A Siemens MAGNETOM Prisma 3T MRI clinical scanner (Siemens Healthcare Erlangen, Germany) and 16-channel phased-array paediatric head and neck coil were used to acquire 3D T2-weighted SPACE images (T2w) (voxel size = 1 mm isotropic, TE = 409 ms and TR = 3200 ms) and axial dMRI data. Diffusion MRI (dMRI) images were acquired in two separate acquisitions to reduce the time needed to re-acquire any data lost to motion artifacts: the first acquisition consisted of 8 baseline volumes (b = 0 s/mm^2^ [b0]) and 64 volumes with b = 750 s/mm^2^, the second consisted of 8 b0, 3 volumes with b = 200 s/mm^2^, 6 volumes with b = 500 s/mm^2^ and 64 volumes with b = 2500 s/mm^2^. An optimal angular coverage for the sampling scheme was applied. In addition, an acquisition of 3 b0 volumes with an inverse phase encoding direction was performed. All dMRI images were acquired using single-shot spin-echo echo planar imaging (EPI) with 2-fold simultaneous multislice and 2-fold in-plane parallel imaging acceleration and 2-mm isotropic voxels; all three diffusion acquisitions had the same parameters (TR/TE 3500/78.0 ms).

MRI scans were acquired at term equivalent age. Infants were scanned in natural sleep with monitoring of pulse oximetry, electrocardiography and temperature. Flexible ear plugs and neonatal earmuffs (MiniMuffs Natus Medical Inc., CA) were used for acoustic protection. All scans were supervised by a doctor or nurse trained in neonatal resuscitation. Conventional images were reported by an experienced paediatric radiologist (A.J.Q.) using a structured system.^23^ Images with evidence of focal parenchymal injury (porencephalic cyst or cystic periventricular leukomalacia) or post-haemorrhagic ventricular dilatation were excluded from image analysis.

### Image processing

Diffusion MRI processing was performed as follows: for each subject the two dMRI acquisitions were first concatenated and then denoised using a Marchenko–Pastur-PCA-based algorithm; the eddy current, head movement, and EPI geometric distortions were corrected using outlier replacement and slice-to-volume registration; bias field inhomogeneity correction was performed by calculating the bias field of the mean b0 volume and applying the correction to all the volumes.^24^

The T2w images were processed using the minimal processing pipeline of the developing human connectome project (dHCP) allowing surface reconstruction from the tissue segmentation. Bias field corrected T2w, the brain masks, the tissue segmentation, the label parcellation and the surfaces reconstruction were obtained.^25^ Finally, the T2w images were co-registered to the mean b0 using boundary-based registration.

### Data quality control

Raw structural and diffusion images were visually inspected. In addition, eddy QC was used to quantify the absolute and relative motion displacement across all volumes for each subject.^26^ The distribution of absolute and relative motion was then compared across all three groups: preterm infants with low breast milk exposure, preterm infants with high breast milk exposure and term controls. There were no significant group wise differences in absolute or relative motion (Figure 1).

**Figure 1.**
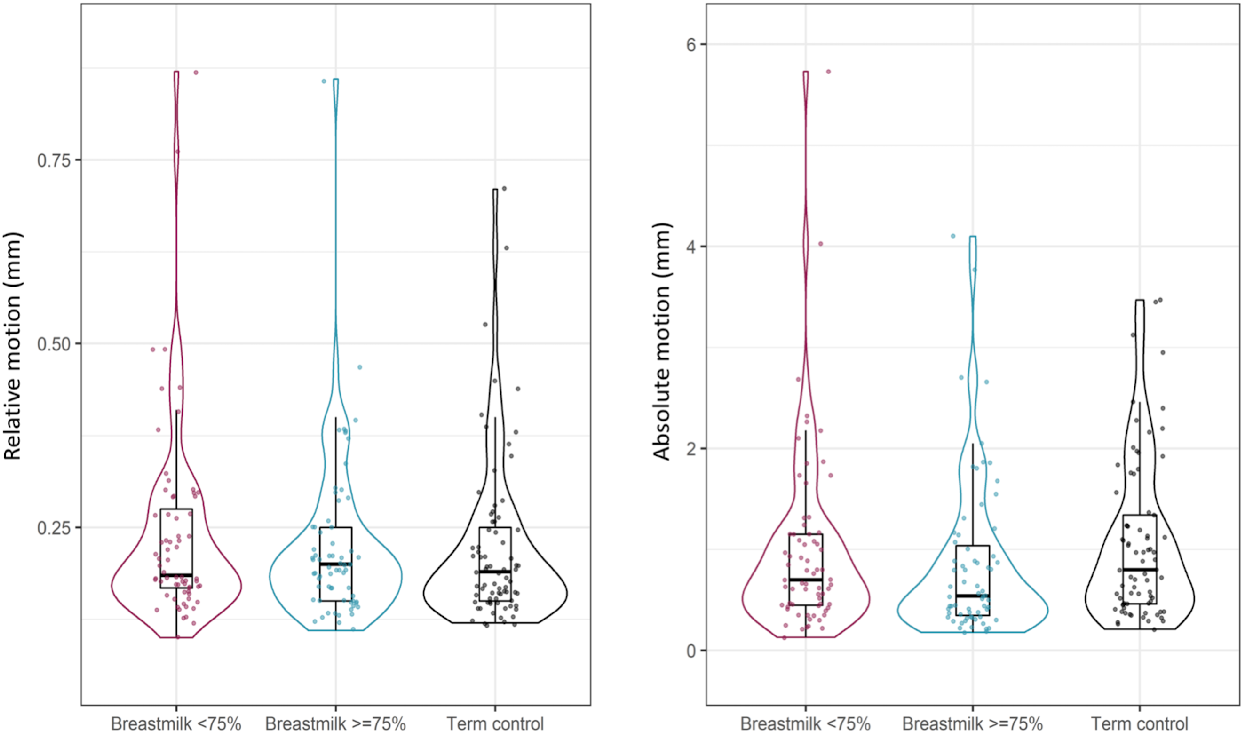
Data quality control. Violin and box plots demonstrating (A) relative and (B) absolute motion in preterm infants with low breast milk exposure (shown in magenta), preterm infants with high breast milk exposure (shown in blue) and term-born controls (shown in black).

### Structural analysis

The volumes were calculated from the tissue and parcellation obtained from the dHCP pipeline.^25^ The area of interest in this study was the cortical gray matter (cGM). The cGM volume was normalised using the total tissue volume (TTV), which included all brain gray matter and white matter tissue. In addition to the volumes (cGM and TTV), we calculated the gyrification index, thickness, sulcal depth, curvature and surface area. All metrics were obtained using the scripts provided in https://github.com/amakropoulos/structural-pipeline-measures.

### Microstructural analysis

From the dMRI, the diffusion tensor (DTI) maps and Neurite Orientation and Dispersion Imaging (NODDI) maps^27^ were generated. For the DTI maps (fractional anisotropy [FA], mean diffusivity [MD], axial diffusivity [AD] and radial diffusivity [RD]) only the b = 750 s/mm^2^ shell was used. The NODDI maps (neurite density index [NDI], isotropic volume fraction [ISO] and orientation dispersion index [ODI]) were calculated using the recommended value for the parallel intrinsic diffusivity of neonatal gray matter (1.25 μm^2^/m).^28^

The mask of the cGM was propagated to the dMRI using the previously computed transformation. To reduce partial volume effects due to CSF contamination, voxels with ISO > 0.5 were excluded from the cGM mask. Finally, the mean value for each of the diffusion derived maps was calculated for the cGM. Representative brain maps are shown in Figure 2.

**Figure 2.**
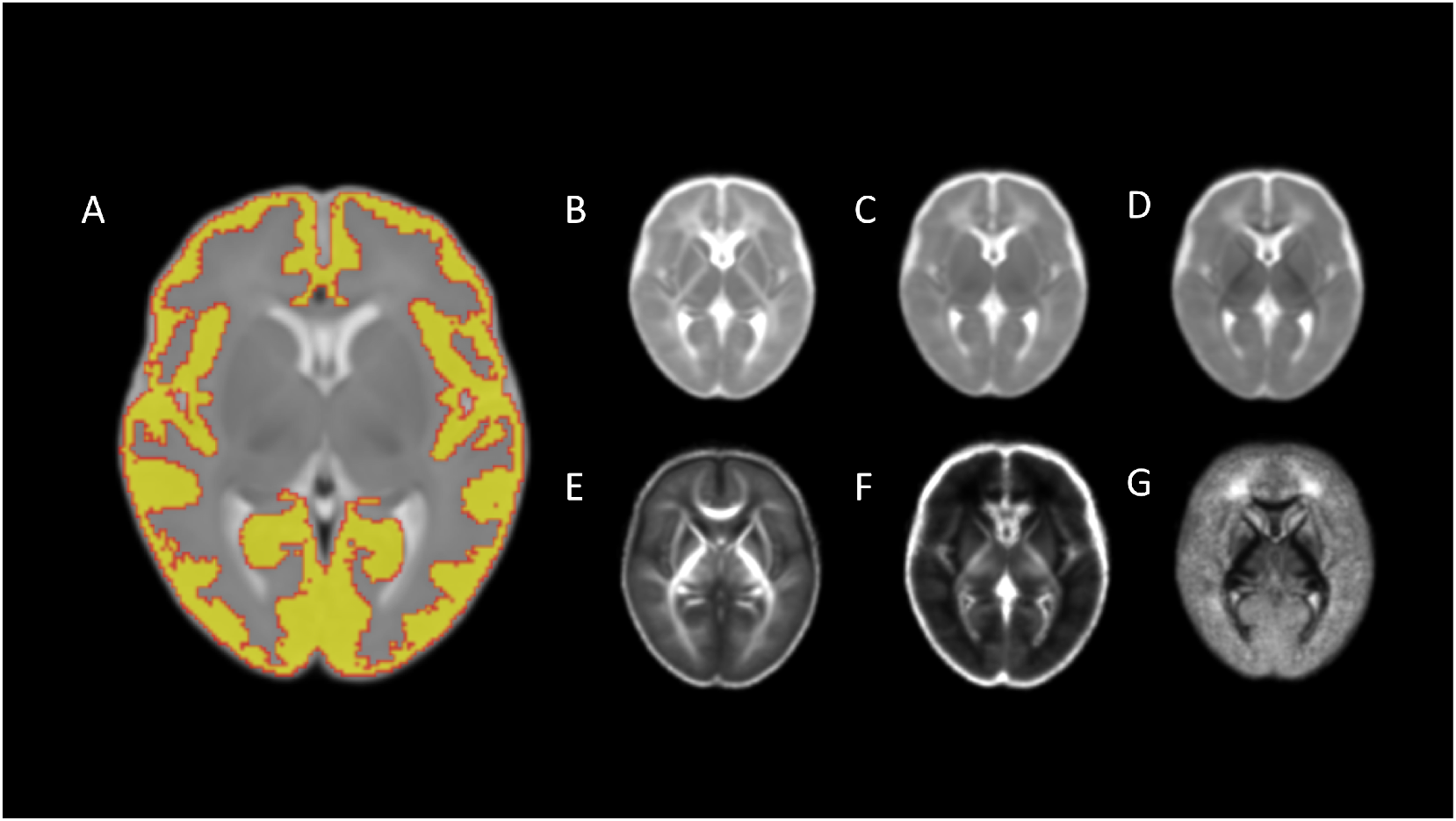
Representative brain maps. Averaged maps for (A) cortical segmentation, (B) axial diffusivity, (C) mean diffusivity, (D) radial diffusivity, (E) fractional anisotropy, (F) neurite density index, (G) orientation dispersion index.

### Statistical analysis

For clinical and demographic features of study participants, categorical variables are shown in absolute numbers with percentages (%), continuous, normally distributed variables as means with standard deviations (SD) and continuous, non-normally distributed variables as medians with ranges. One-way ANOVA was used to compare the means of normally distributed continuous variables between the three groups, Kruskal-Wallis rank sum test was applied to compare medians of non-normally distributed continuous variables and Fischer’s exact test was used to test for significant differences in categorical variables. In two group comparisons of preterm infants with low breast milk exposure versus high breast milk exposure, two-sample t-tests were used to compare the means of normally distributed continuous variables, Wilcoxon rank-sum tests were applied to compare medians of non-normally distributed continuous variables and Fischer’s exact test was used to test for significant differences in categorical variables.

For three group analyses comparing preterm infants with low breast milk exposure, preterm infants with high breast milk exposure and term controls, brain features of interest were scaled (z-transformed) and adjusted for age at MRI. One-way ANOVA was performed for three group comparisons. Normal distribution of the model residuals was tested with the Shapiro-Wilk test; homogeneity of variances was tested using the Levene test. When the assumption for homogeneity of variances was not met, Welch ANOVA was performed. When the assumption for normal distribution of the residuals was not met, Kruskal-Wallis test was performed as the nonparametric alternative. Pairwise Student T-test or Mann-Whitney U-test were performed as post-hoc tests.

For cortical brain imaging features which demonstrated a relationship with breast milk exposure in the three group comparison, we then performed a comparison of preterm infants with low breast milk exposure versus high breast milk exposure, in an additional model with adjustment for gestational age (GA) at birth, age at MRI and bronchopulmonary dysplasia (BPD). Adjustments were made by fitting a linear model of each z-transformed brain feature on the set of covariates and retaining the residuals. Two group comparisons were performed using two-sample T-tests for normally distributed variables and Mann-Whitney U-tests for variables that did not have a normal distribution.

Reported *p*-values were adjusted for false-discovery rate using the Benjamini-Hochberg procedure within each experiment and effect sizes were calculated using Cohen’s *d*. All statistical analyses were performed in R (version 4.05).

### Data availability

Requests for original image data will be considered through the BRAINS governance process (www.brainsimagebank.ac.uk), and all data supporting the findings of the study are available from the corresponding author, upon reasonable request.

## Results

### Participant characteristics

212 infants were studied: 135 preterm infants (mean GA 30^+2^ weeks, range 22^+1^ to 32^+6^) and 77 term-born infants (mean GA 39^+4^ weeks, range 36^+3^ to 42^+1^). Within the preterm group, 68 received exclusive breast milk feeds for <75% of neonatal inpatient days and 67 received exclusive breast milk for ≥75% of neonatal inpatient days. The median proportion of inpatient days preterm infants received exclusive breast milk feeds was 74%. Clinical and demographic features of participants are detailed in Table 1. Within the preterm breast milk groups, there were no statistically significant differences in GA at birth, birthweight z-score or sex. However, age at scan and the prevalence of BPD differed between groups so these were included as covariates in subsequent analyses.

**Table 1.**
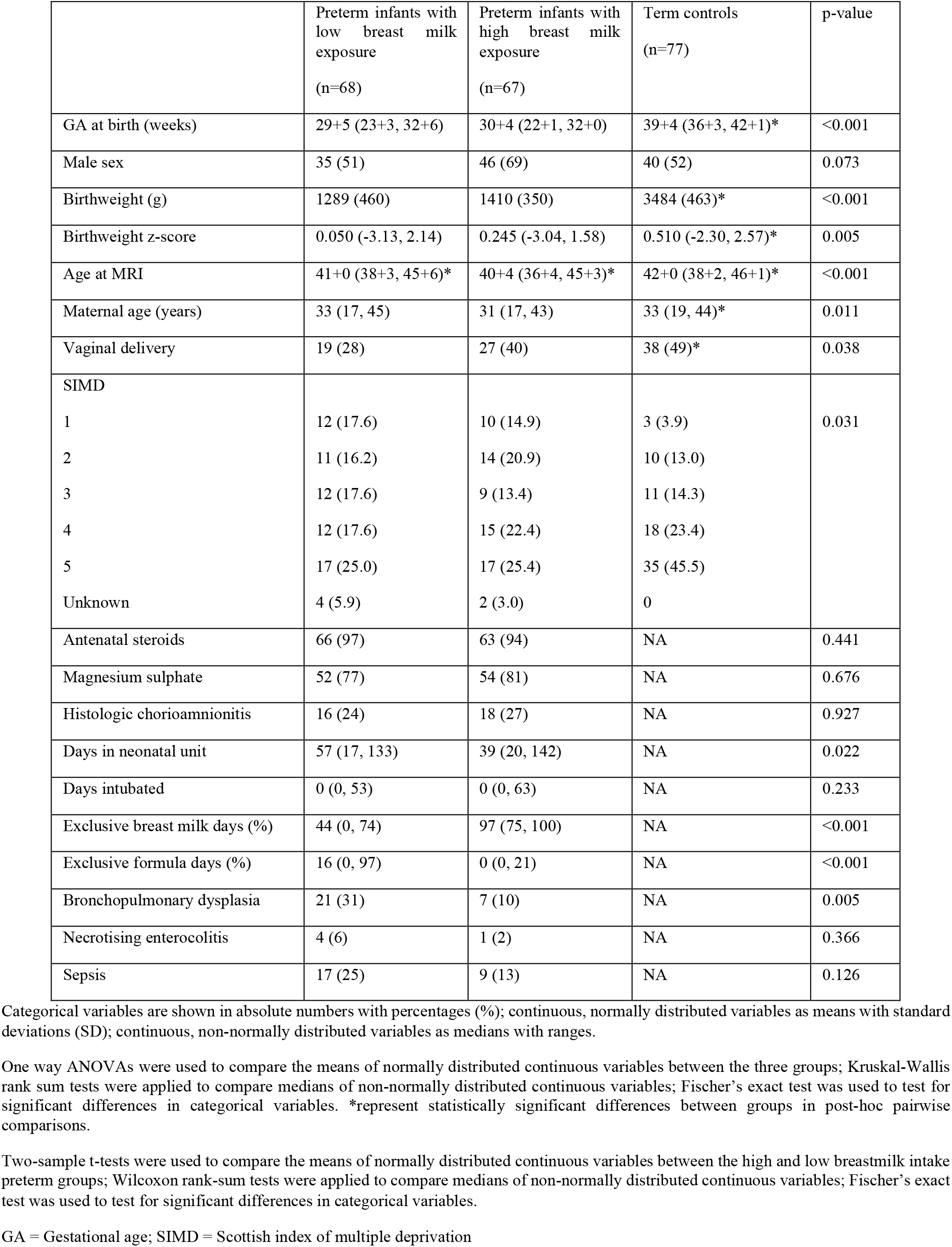
Clinical and demographic characteristics of participants.

### Breast milk exposure and cortical macrostructure

After adjustment for total tissue volume (TTV), preterm infants who received exclusive breast milk feeds for ≥75% of inpatient days had a lower relative cortical gray matter volume (*d*=0.47, *p*=0.014) and reduced cortical thickness (*d*=0.42, *p*=0.039) compared to preterm infants who received exclusive breast milk feeds for <75% of inpatient days (Figure 3B and 3D). There were no significant differences in relative cortical gray matter volume (*d*=0.04, *p*=0.997) or cortical thickness (*d*=0.11, *p*=0.437) between preterm infants who received exclusive breast milk feeds for ≥75% of inpatient days and term-born controls.

**Figure 3.**
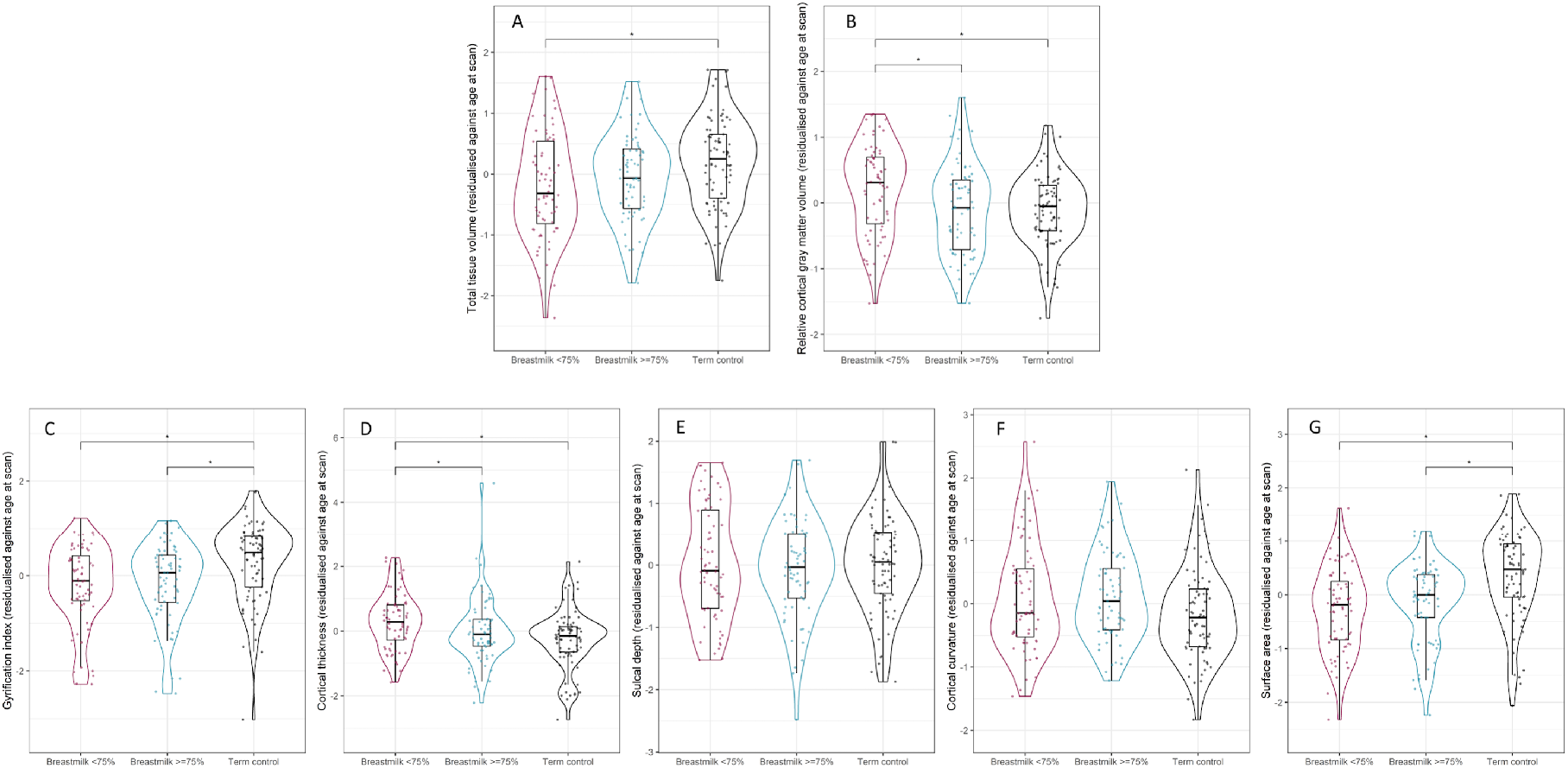
Breast milk exposure and cortical structure. Violin and box plots showing (A) total tissue volume, (B) relative cortical gray matter volume, (C) gyrification index, (D) cortical thickness, (E) sulcal depth, (F) cortical curvature and (G) surface area in preterm infants with low breast milk exposure (shown in magenta), preterm infants with high breast milk exposure (shown in blue) and term-born controls (shown in black). *represents statistically significant pairwise comparisons after FDR correction.

The gyrification index (*d*=0.74 *p*=0.001) and surface area (*d*=0.61, *p*<0.001) were both higher in term infants compared to preterm infants with high breast milk exposure (Figure 3C and 3E), but within the preterm group there was no effect of breast milk exposure on these features. There were no group differences in sulcal depth or cortical curvature.

In the fully adjusted model contrasting the two preterm groups, relative cortical gray matter volume remained significantly lower in preterm infants who received exclusive breast milk feeds for ≥75% of inpatient days when compared to those who received exclusive breast milk feeds for <75% of inpatient days after adjustment for GA at birth, age at MRI and BPD (*d*=0.53, *p*=0.021). Cortical thickness was lower in preterm infants who received exclusive breast milk feeds for ≥75% of inpatient days, but the difference was not statistically significant following correction for multiple tests (*d=*0.34 *p=*0.06).

### Breast milk exposure and cortical microstructure

Preterm infants who received exclusive breast milk feeds for ≥75% of inpatient days had higher mean cortical FA (*d*=0.38, *p*=0.037) and lower mean cortical RD (*d*=0.38, *p*=0.039) compared to preterm infants who received exclusive breast milk feeds for <75% of inpatient days (Figure 4A and 4D).

**Figure 4.**
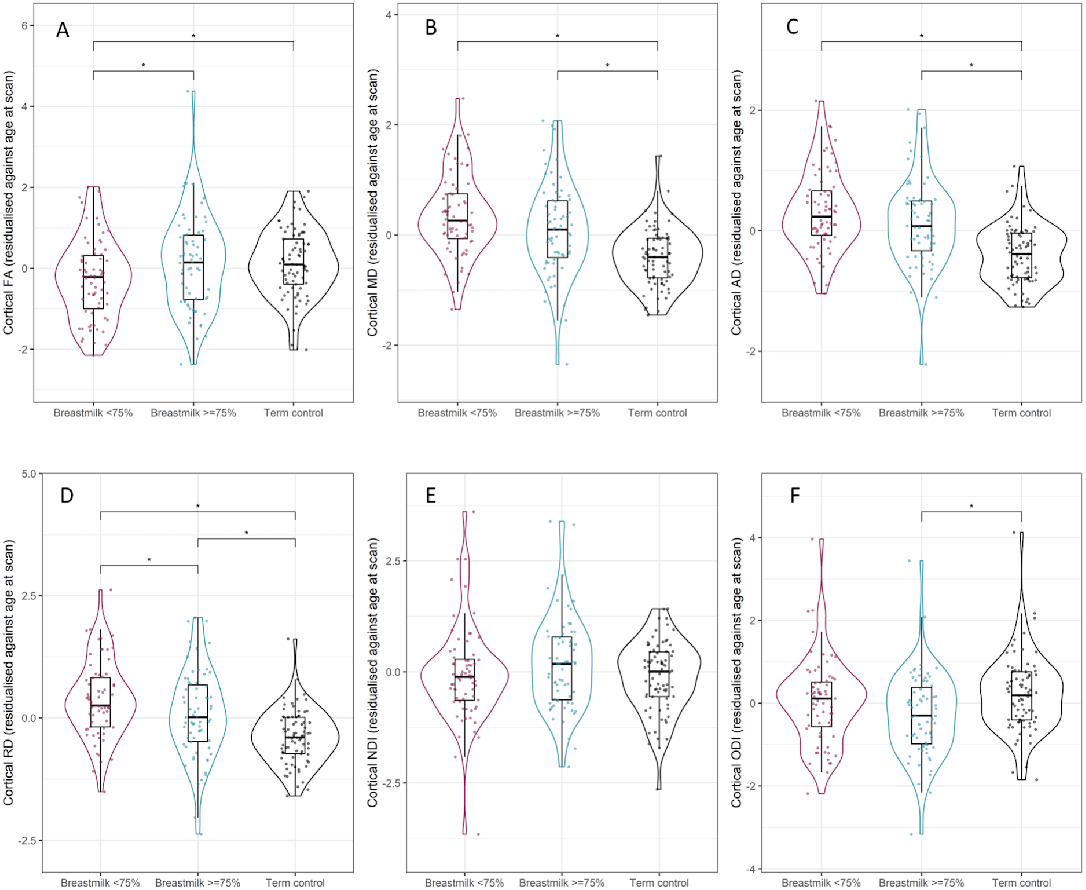
Breast milk exposure and cortical microstructure. Violin and box plots showing mean cortical (A) fractional anisotropy, (B) mean diffusivity, (C) radial diffusivity, (D) axial diffusivity, (E) neurite density index and (F) orientation dispersion index in preterm infants with low breast milk exposure (shown in magenta), preterm infants with high breast milk exposure (shown in blue) and term-born controls (shown in black). *represents statistically significant pairwise comparisons after FDR correction.

There were significant differences in mean cortical RD across all three groups but effect sizes demonstrated a greater difference between preterm infants with low breast milk exposure and term-born controls (*d*=1.1, *p*<0.001) compared to preterm infants with high breast milk exposure versus term-born controls (*d*=0.8, *p*<0.001). The mean cortical FA of preterm infants who received exclusive breast milk feeds for ≥75% of inpatient days was similar to that of term-born controls (*d*=0.03, *p*=0.918).

Mean cortical MD (*d*=0.64, *p*<0.001) and AD (*d*=0.84, *p*<0.001) were lower (Figure 4B and 4C) and cortical ODI (*d*=0.45, *p*=0.005) was higher (Figure 4F) in term infants compared to preterm infants with high breast milk exposure but within the preterm group there was no effect of breast milk exposure on these features. There were no group differences in NDI.

In the fully adjusted model comparing the preterm groups, there were no significant differences in the water diffusion parameters of preterm infants in the high versus low breast milk exposed groups after adjustment for GA at birth, age at MRI and BPD. Preterm infants who received exclusive breast milk feeds for ≥75% of inpatient days had higher FA and lower RD but statistical significance did not survive correction for multiple tests (FA *d*=0.4 *p*=0.05, RD *d*=0.33 *p*=0.06).

## Discussion

The weeks following preterm birth represent a critical window of rapid cortical growth and development which is fundamental to the structural and functional organisation of the brain. By combining nutritional data with brain MRI, we show that high compared to low exposure to exclusive breast milk feeds during this period is associated with lower cortical gray matter volume, thickness and radial diffusivity, and higher fractional anisotropy at term-equivalent age. These findings suggest that higher exposure to breast milk in early life is associated with improved cortical maturation in preterm infants resulting in an imaging phenotype that more closely resembles that of infants born at term.

Preterm birth is associated with a thicker cortex at term-equivalent age when compared to term-born controls^29^ and an atypical trajectory of cortical thinning is suggested by studies of preterm born children^30, 31^ and adolescents.^32, 33^ Healthy cortical growth, expansion and connectivity is critical to the emergence of cognitive functions. Intellectual ability in childhood is associated with a thinner cortex^34^ and increased cortical thickness is associated with neurodevelopmental conditions such as autism spectrum disorder.^35^

Highly regulated spatiotemporal gene expression is crucial for the cellular composition and cytoarchitecture of the developing neocortex and the timing of progenitor cell expansion is a key determinant of cortical size.^36^ Cortical thickness is determined by the number of neuronal cells within cortical columns, which are generated by transient amplifying intermediate progenitor cells and divide symmetrically at the ventricular surface within a radial unit before migrating to their final location within the cortex during development.^37^ Cortical overgrowth may therefore represent increased neurogenesis due to altered proliferation kinetics of intermediate progenitor cells, or altered migration of neurons to the appropriate layers of the developing neocortex. Altered cortical progenitor proliferation has been observed in 3D organoid models of preterm brain injury^38^ and altered migration of interneurons into the developing cortex is a recognised feature of brain dysmaturation after preterm birth.^39^ Our data suggest that higher breast milk exposure following preterm birth improves cortical organisation, offering a potential mechanism underlying the beneficial effects of breast milk exposure on cognitive outcomes.

Early in development, cortical microstructure is characterised by anisotropic water diffusion due to the simple cellular processes of radial glia and apical dendrites which are radially oriented. As the cortex matures, dendritic arborisation is characterised by a reduction in FA due to the geometrical dispersion of processes, followed by a subsequent increase due to neuronal density and organisation.^40^ We found that preterm infants with high breast milk exposure and term-born controls had higher mean cortical FA when compared to preterm infants with low breast milk exposure, suggesting that breast milk exposure is associated with microstructural maturation of the developing cortex at term-equivalent age. Our findings are consistent with recent work demonstrating higher cortical FA at two months of age in breastfed rhesus macaques when compared to those fed formula milk.^41^

Breast milk contains a variety of constituents that may support healthy cortical development including optimal protein and lipid composition, micronutrient availability and non-nutritive factors such as immunoglobulin, lactoferrin, lysozyme, human milk oligosaccharides (HMOs) and microRNAs (miRNAs) with beneficial effects on immune system development and establishment of the gut microbiome.^42^ Previous work has shown that the cognitive benefits of breast milk exposure following preterm birth may be mediated by a reduction in inflammation due to infection or necrotising enterocolitis.^3^ The beneficial effect of breast milk on cortical development may be mediated by immunomodulation through several different mechanisms. Breast milk is rich in miRNAs: small non-coding RNAs that regulate gene expression and protein synthesis at the post-transcription level. Breast milk profiling has identified several miRNAs with potential to modulate early life immune function, lipid metabolism and developmental programming.^43^ miRNAs have been implicated in the regulation of neuroinflammation^44^ and cortical development, influencing neural stem cell expansion and fate specification.^45, 46^

Breast milk feeding is a key determinant of early life gut microbial diversity, composition, and maturity.^47^ The high abundance of HMOs in breast milk provide a nutrient source for Bifidobacterium species, which predominate the gut microbiome of breastfed infants and are integral to early life immune development and regulation.^48^ Studies have described associations between the infant gut microbiome and neurocognitive outcomes in childhood following preterm birth^49^ and in healthy term-born controls^50^ but further work is needed to identify the underlying neurobiological mechanisms.

To our knowledge, this is the first study to examine the effect of early breast milk exposure on cortical development in very preterm infants. Strengths of the study include: (1) Prospective study design with a robust method to reliably ascertain breast milk exposure, (2) A comprehensive assessment of cortical imaging features including volume, expansion, folding and microstructure using diffusion tensor imaging and biophysical modelling, (3) The inclusion of term-born infants to demonstrate imaging features of healthy cortical maturation, and (4) The timing of brain MRI at term-equivalent age to exclude potential confounding by infant nutrition and determinants of brain development operating in the home and family environment following discharge from the neonatal unit.

A limitation of this work is that we were unable to study effects of donor versus maternal expressed breast milk and sample size was insufficient to investigate the effect of sex or socioeconomic status on cortical development. However, our study population showed no sex preponderance and included families from all 5 quintiles of the Scottish Index of Multiple Deprivation. The preterm groups were balanced for GA at birth, exposure to antenatal steroids and magnesium sulphate, and several co-morbidities of preterm birth, but we cannot exclude the possibility that other unknown confounders may contribute to the differences we observed. The main limitation of imaging the developing cortex is image resolution and cortical feature estimates may be biased by partial volume averaging with neighbouring tissue or cerebrospinal fluid or impacted by motion artefact; notably, the data quality control procedure revealed no difference in absolute or relative head motion between groups. Furthermore, we observed differences in the cortical gyrification index, surface area, MD and ODI in preterm infants versus term-born controls consistent with previous reports.^29^

We chose to study the effect of breast milk exposure on whole cortex measures in light of previous work demonstrating global effects of nutrition on the developing brain.^4, 5, 8, 19, 20^ However, regional differences in maturation have been described and differential effects of breast milk according to cortical network would be interesting to consider in future work. Finally, the cross-sectional nature of our data meant that we could not study the effect of breast milk feeding on cortical development beyond the neonatal period. Longitudinal studies are needed to investigate whether the association between breast milk exposure and cortical maturation persists beyond term-equivalent age and to assess the functional consequences of these observations.

## Conclusion

High versus low breast milk exposure in the weeks following preterm birth is associated with imaging features that more closely resemble the cortical maturation of healthy infants born at full term. Healthy development of the cerebral cortex is essential for the establishment of neural networks which are foundational for the emergence of cognition. Breast milk may offer an intervention to optimise early cortical development following preterm birth, thereby reducing the risk of later neurocognitive impairment and psychiatric disease.

## Data Availability

All data produced in the present study are available upon reasonable request to the authors

## Acknowledgements

We are grateful to the families who consented to take part in the study and to the radiographers at the Edinburgh Imaging Facility at the Royal Infirmary of Edinburgh for infant scanning.

## Funding

This work was supported by Theirworld (www.theirworld.org) and was undertaken in the MRC Centre for Reproductive Health, which is funded by MRC Centre Grant (MRC G1002033). KV is funded by the Wellcome Translational Neuroscience PhD Programme at the University of Edinburgh (108890/Z/15/Z). PG is partly supported by the Wellcome-University of Edinburgh ISSF3 (IS3-R1.1320/21). MJT is supported by NHS Lothian Research and Development Office.

## Conflicts of Interest

The authors have nothing to report.

## Notes

### Competing Interest Statement

The authors have declared no competing interest.

### Funding Statement

The study was funded by Theirworld (registered charity 1092312

### Author Declarations

National Research Ethics Service (South East Scotland Committee) granted ethical approval.

## References

1. Vohr BR, Poindexter BB, Dusick AM, et al. Persistent beneficial effects of breast milk ingested in the neonatal intensive care unit on outcomes of extremely low birth weight infants at 30 months of age. Pediatrics. 2007 Oct;120(4):e953–9.

2. Belfort MB, Anderson PJ, Nowak VA, et al. Breast Milk Feeding, Brain Development, and Neurocognitive Outcomes: A 7-Year Longitudinal Study in Infants Born at Less Than 30 Weeks’ Gestation. The Journal of pediatrics. 2016 Oct;177:133-9.e1.

3. Lapidaire W, Lucas A, Clayden JD, Clark C, Fewtrell MS. Human milk feeding and cognitive outcome in preterm infants: the role of infection and NEC reduction. Pediatric research. 2021 2021/06/24.

4. Blesa M, Sullivan G, Anblagan D, et al. Early breast milk exposure modifies brain connectivity in preterm infants. NeuroImage. 2019 Jan 1;184:431–9.

5. Ottolini KM, Andescavage N, Kapse K, Jacobs M, Limperopoulos C. Improved brain growth and microstructural development in breast milk–fed very low birth weight premature infants. Acta Paediatrica. 2020;109(8):1580–7.

6. Isaacs EB, Fischl BR, Quinn BT, Chong WK, Gadian DG, Lucas A. Impact of breast milk on intelligence quotient, brain size, and white matter development. Pediatric research. 2010 Apr;67(4):357–62.

7. Ou X, Andres A, Pivik RT, et al. Voxel-Based Morphometry and fMRI Revealed Differences in Brain Gray Matter in Breastfed and Milk Formula-Fed Children. AJNR American journal of neuroradiology. 2016 Apr;37(4):713–9.

8. Luby JL, Belden AC, Whalen D, Harms MP, Barch DM. Breastfeeding and Childhood IQ: The Mediating Role of Gray Matter Volume. Journal of the American Academy of Child and Adolescent Psychiatry. 2016 May;55(5):367–75.

9. Kafouri S, Kramer M, Leonard G, et al. Breastfeeding and brain structure in adolescence. International Journal of Epidemiology. 2012;42(1):150–9.

10. Kostović I, Sedmak G, Judaš M. Neural histology and neurogenesis of the human fetal and infant brain. NeuroImage. 2019 Mar;188:743–73.

11. Volpe JJ. Dysmaturation of Premature Brain: Importance, Cellular Mechanisms, and Potential Interventions. Pediatric neurology. 2019 Jun;95:42–66.

12. Ball G, Srinivasan L, Aljabar P, et al. Development of cortical microstructure in the preterm human brain. Proceedings of the National Academy of Sciences of the United States of America. 2013 Jun 4;110(23):9541–6.

13. Shimony JS, Smyser CD, Wideman G, et al. Comparison of cortical folding measures for evaluation of developing human brain. NeuroImage. 2016 Jan 15;125:780–90.

14. Zhang Y, Inder TE, Neil JJ, et al. Cortical structural abnormalities in very preterm children at 7 years of age. NeuroImage. 2015 Apr 1;109:469–79.

15. Barnes-Davis ME, Williamson BJ, Merhar SL, Holland SK, Kadis DS. Extremely preterm children exhibit altered cortical thickness in language areas. Scientific reports. 2020 2020/07/02;10(1):10824.

16. Nam KW, Castellanos N, Simmons A, et al. Alterations in cortical thickness development in preterm-born individuals: Implications for high-order cognitive functions. NeuroImage. 2015 Jul 15;115:64–75.

17. Keunen K, Išgum I, van Kooij BJ, et al. Brain Volumes at Term-Equivalent Age in Preterm Infants: Imaging Biomarkers for Neurodevelopmental Outcome through Early School Age. The Journal of pediatrics. 2016 May;172:88–95.

18. Kline JE, Illapani VSP, He L, Altaye M, Logan JW, Parikh NA. Early cortical maturation predicts neurodevelopment in very preterm infants. Archives of disease in childhood Fetal and neonatal edition. 2019 Nov 8.

19. Schneider J, Fischer Fumeaux CJ, Duerden EG, et al. Nutrient Intake in the First Two Weeks of Life and Brain Growth in Preterm Neonates. Pediatrics. 2018 Mar;141(3).

20. Coviello C, Keunen K, Kersbergen KJ, et al. Effects of early nutrition and growth on brain volumes, white matter microstructure, and neurodevelopmental outcome in preterm newborns. Pediatric research. 2018 Jan;83(1-1):102–10.

21. Rozé JC, Darmaun D, Boquien CY, et al. The apparent breastfeeding paradox in very preterm infants: relationship between breast feeding, early weight gain and neurodevelopment based on results from two cohorts, EPIPAGE and LIFT. BMJ Open. 2012;2(2):e000834.

22. Boardman JP, Hall J, Thrippleton MJ, et al. Impact of preterm birth on brain development and long-term outcome: protocol for a cohort study in Scotland. BMJ Open. 2020;10(3):e035854.

23. Leuchter RH, Gui L, Poncet A, et al. Association between early administration of high-dose erythropoietin in preterm infants and brain MRI abnormality at term-equivalent age. Jama. 2014 Aug 27;312(8):817–24.

24. Blesa M, Galdi P, Cox SR, et al. Hierarchical Complexity of the Macro-Scale Neonatal Brain. Cerebral Cortex. 2020.

25. Makropoulos A, Counsell SJ, Rueckert D. A review on automatic fetal and neonatal brain MRI segmentation. NeuroImage. 2018 Apr 15;170:231–48.

26. Bastiani M, Cottaar M, Fitzgibbon SP, et al. Automated quality control for within and between studies diffusion MRI data using a non-parametric framework for movement and distortion correction. NeuroImage. 2019 2019/01/01/;184:801–12.

27. Zhang H, Schneider T, Wheeler-Kingshott CA, Alexander DC. NODDI: practical in vivo neurite orientation dispersion and density imaging of the human brain. NeuroImage. 2012 Jul 16;61(4):1000–16.

28. Guerrero JM, Adluru N, Bendlin BB, et al. Optimizing the intrinsic parallel diffusivity in NODDI: An extensive empirical evaluation. PloS one. 2019;14(9):e0217118.

29. Dimitrova R, Pietsch M, Ciarrusta J, et al. Preterm birth alters the development of cortical microstructure and morphology at term-equivalent age. NeuroImage. 2021 2021/11/01/;243:118488.

30. Mürner-Lavanchy I, Steinlin M, Nelle M, et al. Delay of cortical thinning in very preterm born children. Early human development. 2014 Sep;90(9):443–50.

31. Vandewouw MM, Young JM, Mossad SI, et al. Mapping the neuroanatomical impact of very preterm birth across childhood. Human brain mapping. 2020 Mar;41(4):892–905.

32. Nagy Z, Lagercrantz H, Hutton C. Effects of preterm birth on cortical thickness measured in adolescence. Cerebral cortex (New York, NY : 1991). 2011 Feb;21(2):300–6.

33. Bjuland KJ, Løhaugen GC, Martinussen M, Skranes J. Cortical thickness and cognition in very-low-birth-weight late teenagers. Early human development. 2013 Jun;89(6):371–80.

34. Schnack HG, van Haren NE, Brouwer RM, et al. Changes in thickness and surface area of the human cortex and their relationship with intelligence. Cerebral cortex (New York, NY : 1991). 2015 Jun;25(6):1608–17.

35. Yang DYJ, Beam D, Pelphrey KA, Abdullahi S, Jou RJ. Cortical morphological markers in children with autism: a structural magnetic resonance imaging study of thickness, area, volume, and gyrification. Molecular Autism. 2016 2016/01/25;7(1):11.

36. Nowakowski TJ, Bhaduri A, Pollen AA, et al. Spatiotemporal gene expression trajectories reveal developmental hierarchies of the human cortex. Science (New York, NY). 2017 Dec 8;358(6368):1318–23.

37. Bystron I, Blakemore C, Rakic P. Development of the human cerebral cortex: Boulder Committee revisited. Nature reviews Neuroscience. 2008 Feb;9(2):110–22.

38. Paşca AM, Park JY, Shin HW, et al. Human 3D cellular model of hypoxic brain injury of prematurity. Nature medicine. 2019 May;25(5):784–91.

39. Stolp HB, Fleiss B, Arai Y, et al. Interneuron Development Is Disrupted in Preterm Brains With Diffuse White Matter Injury: Observations in Mouse and Human. Frontiers in physiology. 2019 2019-July-30;10(955).

40. Batalle D, O’Muircheartaigh J, Makropoulos A, et al. Different patterns of cortical maturation before and after 38 weeks gestational age demonstrated by diffusion MRI in vivo. NeuroImage. 2019 Jan 15;185:764–75.

41. Liu Z, Neuringer M, Erdman JW, Jr., et al. The effects of breastfeeding versus formula-feeding on cerebral cortex maturation in infant rhesus macaques. NeuroImage. 2019 Jan 1;184:372–85.

42. Carr LE, Virmani MD, Rosa F, et al. Role of Human Milk Bioactives on Infants’ Gut and Immune Health. Frontiers in immunology. 2021 2021-February-12;12(290).

43. Carrillo-Lozano E, Sebastián-Valles F, Knott-Torcal C. Circulating microRNAs in Breast Milk and Their Potential Impact on the Infant. Nutrients. 2020 Oct 8;12(10).

44. Gaudet AD, Fonken LK, Watkins LR, Nelson RJ, Popovich PG. MicroRNAs: Roles in Regulating Neuroinflammation. The Neuroscientist : a review journal bringing neurobiology, neurology and psychiatry. 2018 Jun;24(3):221–45.

45. Bian S, Hong J, Li Q, et al. MicroRNA cluster miR-17-92 regulates neural stem cell expansion and transition to intermediate progenitors in the developing mouse neocortex. Cell reports. 2013 May 30;3(5):1398–406.

46. Åkerblom M, Jakobsson J. MicroRNAs as Neuronal Fate Determinants. The Neuroscientist : a review journal bringing neurobiology, neurology and psychiatry. 2014 Jun;20(3):235–42.

47. Ho NT, Li F, Lee-Sarwar KA, et al. Meta-analysis of effects of exclusive breastfeeding on infant gut microbiota across populations. Nature communications. 2018 2018/10/09;9(1):4169.

48. Henrick BM, Rodriguez L, Lakshmikanth T, et al. Bifidobacteria-mediated immune system imprinting early in life. Cell. 2021 Jul 22;184(15):3884-98.e11.

49. Seki D, Mayer M, Hausmann B, et al. Aberrant gut-microbiota-immune-brain axis development in premature neonates with brain damage. Cell Host & Microbe. 2021 2021/10/13/;29(10):1558-72.e6.

50. Carlson AL, Xia K, Azcarate-Peril MA, et al. Infant Gut Microbiome Associated With Cognitive Development. Biological psychiatry. 2018 Jan 15;83(2):148–59.

